# Array Comparative Genomic Hybridisation and Droplet Digital PCR uncover recurrent copy number variation of the titin segmental duplication region

**DOI:** 10.1101/2022.03.16.22272470

**Authors:** Lydia Sagath, Vilma-Lotta Lehtokari, Katarina Pelin, Kirsi Kiiski

## Abstract

Intragenic segmental duplication regions are potential hotspots for recurrent copy number variation and possible pathogenic aberrations. Two large sarcomeric genes, nebulin and titin, both contain such segmental duplication regions. Using our custom Comparative Genomic Hybridization array, we have previously shown that a gain or loss of more than one copy of the repeated block of the nebulin triplicate region constitutes a recessive pathogenic mutation. Using targeted array-CGH, similar copy number variants can be detected in the segmental duplication region of titin. Due to the limitations of the array-CGH methodology and the repetitiveness of the region, the exact copy numbers of the blocks could not be determined. Therefore, we developed complementary custom Droplet Digital PCR assays for the titin segmental duplication region to confirm true variation. Our combined methods show that the titin segmental duplication region is subject to recurrent copy number variants, which is surprisingly common. Gains and losses were detected in samples from healthy individuals as well as in samples from patients with different muscle disorders. The copy number variation observed in our cohort is likely benign, but pathogenic copy number variants in the segmental duplication region of titin cannot be excluded. Further investigations are needed, however, this region should no longer be neglected in genetic analyses.

## Introduction

Segmental duplications (SD) are highly identical, 10 – 300 kb long genomic sequences present from two to a few times in the genome, either interspersed or in tandem (1-3). They predispose regions for copy number variants (CNVs) and may thus act as mutational hotspots (4-6).

Titin (*TTN*, MIM ID *188840) is a gargantuan gene highly expressed in skeletal muscle. According to the reference sequence of the longest *TTN* transcript (ENST00000589042.5, CCDS59435.1), it consists of 363 exons. In its middle, it holds a region encoding domains rich in proline (P), glutamate (E), valine (V), and lysine (K), referred to as the PEVK region (7). Within the PEVK region, it withholds a SD region (exons 172–180, 181–189, 190–198, and 203-204). This region consists of a 9-exon-block repeated three times, after which the two first exons of the block appear a fourth time. These two exons are separated from the last exon of the last repeated block (exon 198) by four exons (Fig 1). The first two exons of the block (exons 172-173, 181-182, 190-191, and 203-204, respectively) exist in a ratio of 4:1 compared to the diploid genome, while the respective ratio for the other exons is 3:1. The structure of the *TTN* SD region is depicted in Figure 1.

**Figure 1.**
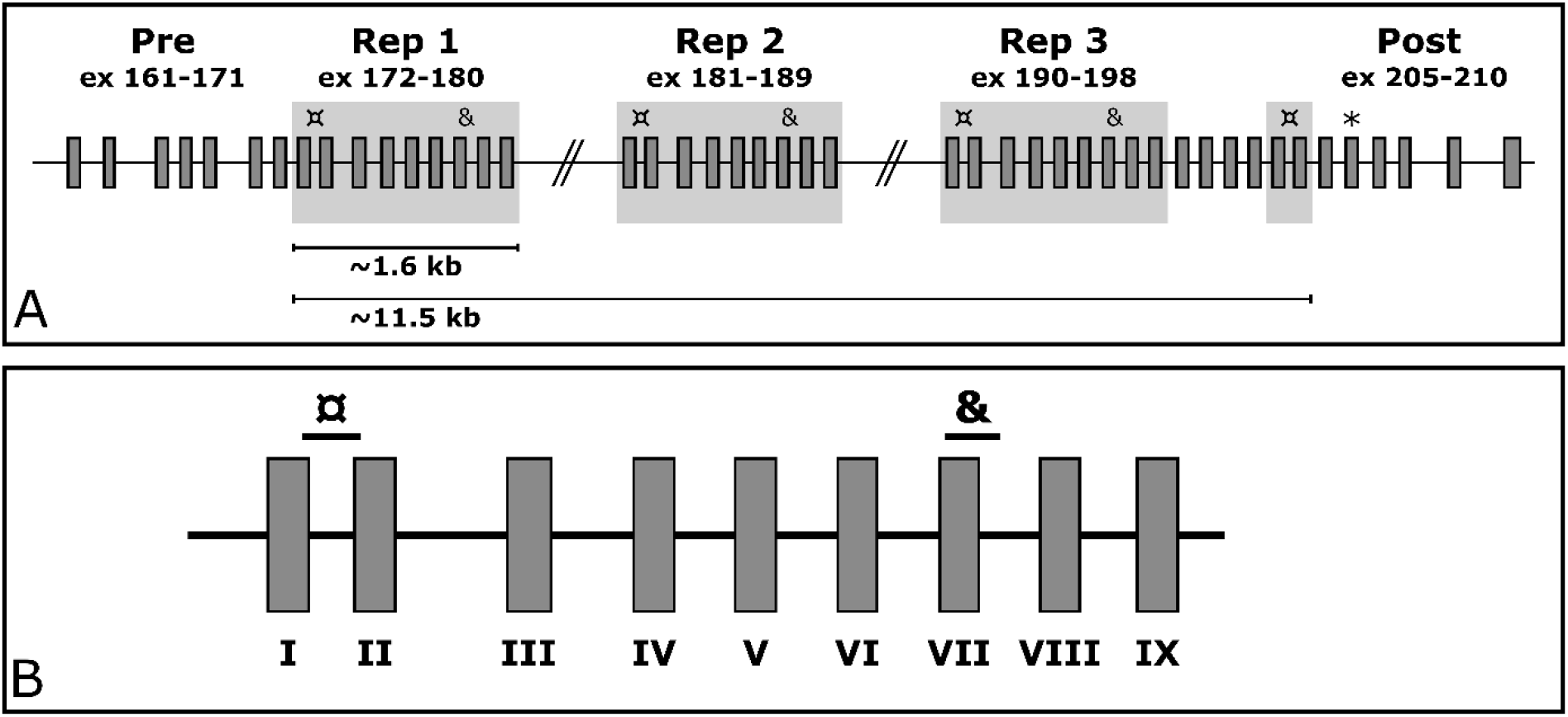
The structure of the *TTN* SD region. Panel A shows the repeated blocks shaded in grey. The custom ddPCR assays targeting the *TTN* SD region exons I and VII are marked with ¤ and & respectively. Panel B shows a zoom-in on the repeated block along with the target locations of the assays. The targeted exons downstream of the segmental duplication are marked with an asterisk (*).

Mutations in *TTN* can cause several different neuromuscular diseases, such as tibial muscular dystrophy and cardiomyopathies, in both recessive and dominant inheritance modes (MIM IDs dilated cardiomyopathy #604145, familial hypertrophic cardiomyopathy #613765, limb-girdle muscular dystrophy type 2J #608807, proximal myopathy #603689, Salih myopathy #611705, and tibial muscular dystrophy #600334) (8-12).

The standard method for routine CNV analysis is still array Comparative Genomic Hybridization (aCGH). However, CNV analysis methods based on massively parallel sequencing data are rapidly improving in accuracy and reliability (13-16). SDs and other repetitive regions still challenge both aCGH and methods based on massively parallel sequencing (MPS) in CNV detection. Designing unique aCGH probes is challenging due to the repetitive nature of these regions, thus these regions are typically avoided in commercial aCGH designs and often left with minimal to no probe coverage. Similarly, the alignment of short sequences challenges the analysis of repetitive regions with MPS-based methods.

We have previously designed and published two validated custom tiling arrays for the detection of CNVs in neuromuscular disorder genes (17, 18). These arrays also cover the *TTN* SD and a similar SD in another muscle gene, the triplicate (TRI) region in nebulin (encoded by the *NEB* gene, MIM ID *161650) to shed light on these regions and their variations (17, 18).

Like *TTN, NEB* is a large structural protein highly expressed in skeletal muscle. Pathogenic variants in *NEB* are a known cause of muscle disorders, such as nemaline myopathy (MIM #256030). Both *TTN* and *NEB* are thought to act as molecular templates, or rulers, for muscle filament length and structure (19-24). As per this Ruler Hypothesis, large enough gains and losses in CN in the *NEB* TRI and *TTN* SD regions may be pathogenic (19-26). It has been shown that gains of two blocks of *NEB* TRI in one allele are, in fact, disease-causing (25).

The major difference between the SD regions of *NEB* and *TTN* is the difference in size – the *NEB* TRI covers altogether 30 kb of genomic region, which is roughly three times more than the *TTN* SD. Its length allows a tiled aCGH design in this region, despite its repetitiveness. The *TTN* SD region only covers around 10 kb in its entirety, severely challenging the tiled probe approach. Furthermore, the *TTN* SD is doubly repetitive; on top of the block repeating, the exons within the block are similar, and share sequences with flanking exons also. Therefore, the resolution and mathematical mode of analysis of aCGH data are challenged by this region, as it heavily relies on the possibility of incorporating suitable probes, and aberration calls are typically made based on several consecutive affected probes.

To allow for large-scale screening of CNVs of the *NEB* TRI region, we have previously developed two custom Droplet Digital PCR (ddPCR) assays targeting the region (27). Here, we present the extension of the study; custom ddPCR assays for the detection of CNVs within the *TTN* SD region. We show, using ddPCR and aCGH data, that the *TTN* SD region is subject to CNVs in a similar fashion to the *NEB* TRI region. To our knowledge, this is the first publication acknowledging CNVs within *TTN* SD to this degree and shows that this region should no longer be neglected in genetic analyses.

## Materials and methods

### Samples

Altogether 62 samples from 42 neuromuscular disorder families were acquired for the study. Of these, 42 were index patient samples, and the remaining 20 samples were from unaffected family members. The patient phenotypes included nemaline myopathy, distal nemaline myopathy, asymmetric distal myopathy, arthrogryposis, and unspecified congenital myopathy.

The DNA stocks had been extracted either from peripheral blood or from saliva, eluted into EDTA, TE-buffer or water, and stored at −20°C. The DNA concentration and quality were checked with DeNovix DS-11 FX+ Spectrophotometer/Fluorometer (DeNovix Inc., Wilmington, DE, USA). Subsequent dilutions for the ddPCR reactions were done in sterile water and stored at 4°C.

### Comparative Genomic Hybridisation array design, protocol, and analysis

All samples were run using aCGH for neuromuscular disorders (NMD-CGH-array) as previously described (18).

The aCGH data were manually aligned for *TTN* and *NEB* to gain a baseline of zero to avoid any subtle differences caused by the genome-wide normalization of the analysis software (CytoSure Interpret Software v.4.11.30, Oxford Gene Technology Ltd, Cambridge, UK). The log2 value for the *TTN* SD region and large regions (124 kb and 157 kb) of the *TTN* gene upstream and downstream of the SD were extracted from the aCGH data. The breakpoints used for normalization of *TTN* aCGH results were Chr2:(179389578_179390615)_(179512818_179513536) (upstream backbone), Chr2:(179533543_179533609)_(179691400_179690759) (downstream backbone), and Chr2:(179518163_179518846)_(179528302_179528492) (segmental duplication region). The breakpoints used for the normalization of *NEB* have been published earlier (27). The genomic locations for the aCGH data are given in the reference genome build Hg19/GRCh37. The normalized log2 value of the *TTN* region was acquired by subtracting the averaged background log2 value from log2 value of the *TTN* SD region.

### Droplet digital PCR

The ddPCR assays were designed, performed, and analysed according to the dMIQE guidelines (28, 29). The dMIQE checklist is available in Supplemental Table 1.

#### Primer and probe design

Custom assays were designed for three regions in *TTN* (NG_011618.3). Two of these target repeated exons within the SD region, and one targets an exon located downstream from the SD region (exon 206, Fig 1A). The assays targeting the SD span from the end of exon 172/181/190/203 to the beginning of exon 173/182/191/204 (from here on referred to as *TTN* SD exon I as per the *TTN* SD exon it begins in) and from the beginning of exon 178/187/196 to the beginning of intron 178/187/196 (from here on referred to as *TTN* SD exon VII) (Fig 1B). The assay targeting the exon downstream of the SD region is from here on referred to as *TTN* Post-SD.

Primers and probes for the assays were designed using Primer3Plus (http://www.bioinformatics.nl/cgi-bin/primer3plus/primer3plus.cgi) (30, 31). Primers were designed to have a melting temperature (Tm) of approximately 60°C, a GC content of 50-60 %, a length of 20 bp, and avoiding putative secondary structures, and G or C repeats over three bases long. The amplicons were not allowed to contain the BsuRI cut site sequence (GGCC) or the EcoRV cut site sequence (GATATC). Amplicon lengths vary from 101 to 120 bp.

Hydrolysis probes were designed to have a Tm of approximately 65°C and a GC content of 30-80%, aiming for a length of 25 bp. Custom probes were labelled with fluorescein amitide (FAM). Tm was calculated by the nearest neighbour method using OligoCalc (32).

An in silico specificity screen was performed using the Standard Nucleotide BLAST blastn suite (https://blast.ncbi.nlm.nih.gov/Blast.cgi), allowing four hits for the *TTN* SD region exon I assay primers and probes, three hits for the *TTN* SD exon VII assay, and one hit for the *TTN* Post-SD assay.

All custom primer and probe sequences along with amplicon lengths and locations within the reference sequences NG_011618.3 and NG_009382.2 are presented in Supplemental Table 2.

The reference used was a commercial *EIF2C1* ddPCR probe assay labelled with hexachloro-fluorescein (HEX) (cat. no. 10031243, Bio-Rad Laboratories Inc.). *EIF2C1*, also known as Argonaute 1 (*AGO1*), is a diploid gene located in the chromosomal region 1p34.3.

The previously published custom ddPCR assay for *NEB* TRI exon VIII (27) was included as a positive control for each sample in every run.

All assays were obtained from Bio-Rad Laboratories, Inc (Hercules, CA, USA) as custom ordered primer/probe assays at a primer:probe ratio of 3.6:1. The concentrations were 900 nM of primer and 250 nM of probe in the final reaction mix.

#### Optimisation of assay conditions

The optimal melting temperature for the assays was determined by running the reactions in a gradient PCR with different melting temperatures. The assays were evaluated using the melting temperatures of 57.5, 58.0, 58.5, 59.0, 59.5, 60.0, 60.5 and 61.0°C. A melting temperature of 59.5°C was chosen as it gave an adequate separation between the droplet clusters in all assays.

#### Assay protocol

The total reaction volume was 20 μl, consisting of 10 ng of genomic template DNA in a volume of 7 μl, 1 μl of custom and reference assay each, 1 μl of restriction enzyme mix, and 10 µl of 2x ddPCR Supermix for Probes (No dUTPs) (cat. no. 1863023, Bio-Rad Laboratories Inc). The restriction enzyme mix contained equal amounts of BsuRI (cat. no. FD0154, Thermo Scientific, Waltham, MA, USA) and EcoRV (cat. no. F0304, Thermo Scientific) for the *TTN* assays, and 1:1 diluted BsuRI in 10x FastDigest Buffer (cat. no. B64, Thermo Scientific) for the *NEB* assay. For each reaction, a total of 22 μl of reaction mix was prepared, of which 20μl was pipetted onto the DG8 Cartridge (cat. no. 1864008, Bio-Rad Laboratories Inc.). The cartridges were covered with DG8 Gaskets (cat. no. 1863009, Bio-Rad Laboratories Inc.).

The reactions were divided into approximately 1 nl droplets using the Droplet Generator QX2000 (Bio-Rad Laboratories Inc.) with Droplet Generator Oil (cat. no. 1863005, Bio-Rad Laboratories Inc.), transferred to ddPCR 96-well plates (cat. no. 12001925, Bio-Rad Laboratories Inc.) by pipetting and sealed with the PX1 PCR Plate Sealer (Bio-Rad Laboratories Inc.). The PCR reaction was performed using the DNA Engine Tetrad 2 Thermal Cycler (Bio-Rad Laboratories Inc.). The cycling steps were 95°C 10 minutes; 40 cycles of (94°C 30 seconds, 59.5°C 1 minute); 98°C 10 minutes; 4°C hold, with a ramp rate of 2°C/sec. The data was then visually inspected on the QuantaSoft Analysis v. 1.7.4.0917 and QuantaSoft Analysis Pro v. 1.0.596.0525 (Bio-Rad Laboratories Inc.) programs.

Each plate contained at least one no-template control for each assay to assess putative contamination. All samples were run in duplicate.

#### Data extraction and filtering

Droplet data was extracted as comma-separated values (CSV) files from the QuantaSoft Analysis software and imported into R-Studio v.1.4.1103 (33, 34).

Subsequent filtering was performed in Microsoft Excel using a minimum threshold of 25 droplets for individual droplet categories and 8,500 accepted droplets per reaction. Data from two successful wells were used in subsequent analyses. In cases of more than two successful wells for the same assay and sample, the two runs with the largest number of accepted droplets were used in the analysis. The data was grouped as normal, gain, and loss samples by the predicted *TTN* SD copy number category based on the respective NMD-CGH-array results.

The range, mean, standard deviation (σ) and coefficient of variation in percent (%CV) were extracted and calculated of the accepted droplet number and target copies per 20 µl, and for the estimated CN by ddPCR and aCGH for the respective regions targeted by the ddPCR assays.

Boxplots were generated using ggplot2 v.3.3.6 (35).

#### Statistical analysis

To assess the performance of the ddPCR assays in CN classification, we performed one-way analysis of variance (ANOVA) tests and post hoc Tukey’s honest significant difference (Tukey’s HSD) test using the ddPCR derived CN and the aCGH-predicted CNV class (either normal, loss, or gain).

A linear regression analysis was performed using the CN estimates from ddPCR and aCGH. In addition, the Pearson correlation coefficient was also calculated from this data. A Pearson correlation coefficient of >0.70 was considered a strong correlation.

To assess reproducibility within experiments, we performed intra-assay analyses separately for all assays using duplicates run within the same experiment.

To assess repeatability between experiments, we performed inter-assay analyses separately for all assays using duplicates run on separate plates.

All statistical analyses were performed in RStudio (v.1.4.1103) using R v.4.0.4. (33, 34).

## Results

### Data overview

Altogether 55 samples passed the initial quality filtering in all assays in at least two parallel wells. Of these, 36 were samples from neuromuscular disorder patients and 19 were samples from unaffected family members. Of all samples, 35 were classed as normal, 11 as losses, and 9 as gains as per the CNV prediction of the NMD-CGH-array for the *TTN* SD region. Gains and losses in the *TTN* SD region were present in both patient and unaffected family member samples. Table 1 shows the number and percentage of samples representing different predicted CN classes of the *TTN* SD. The CNs indicated by ddPCR are shown plotted against the aCGH-determined CN class for each respective targeted region in Figure 2.

**Table 1.** Number and percentage of samples representing different predicted classes of the *TTN* SD according to the NMD-CGH-array. The normal CN for the *NEB* TRI region is 6, and any deviation from this is considered either a loss or a gain.

| *TTN* SD status | Number of samples | % of samples | *NEB* TRI status |  |  |
| --- | --- | --- | --- | --- | --- |
|  |  |  | Loss | Normal | Gain |
| Gain | 9 | 16.4% | 2 | 5 | 2 |
| Normal | 35 | 63.6 % | 1 | 33 | 1 |
| Loss | 11 | 20.0 % | 1 | 9 | 1 |

**Figure 2.**
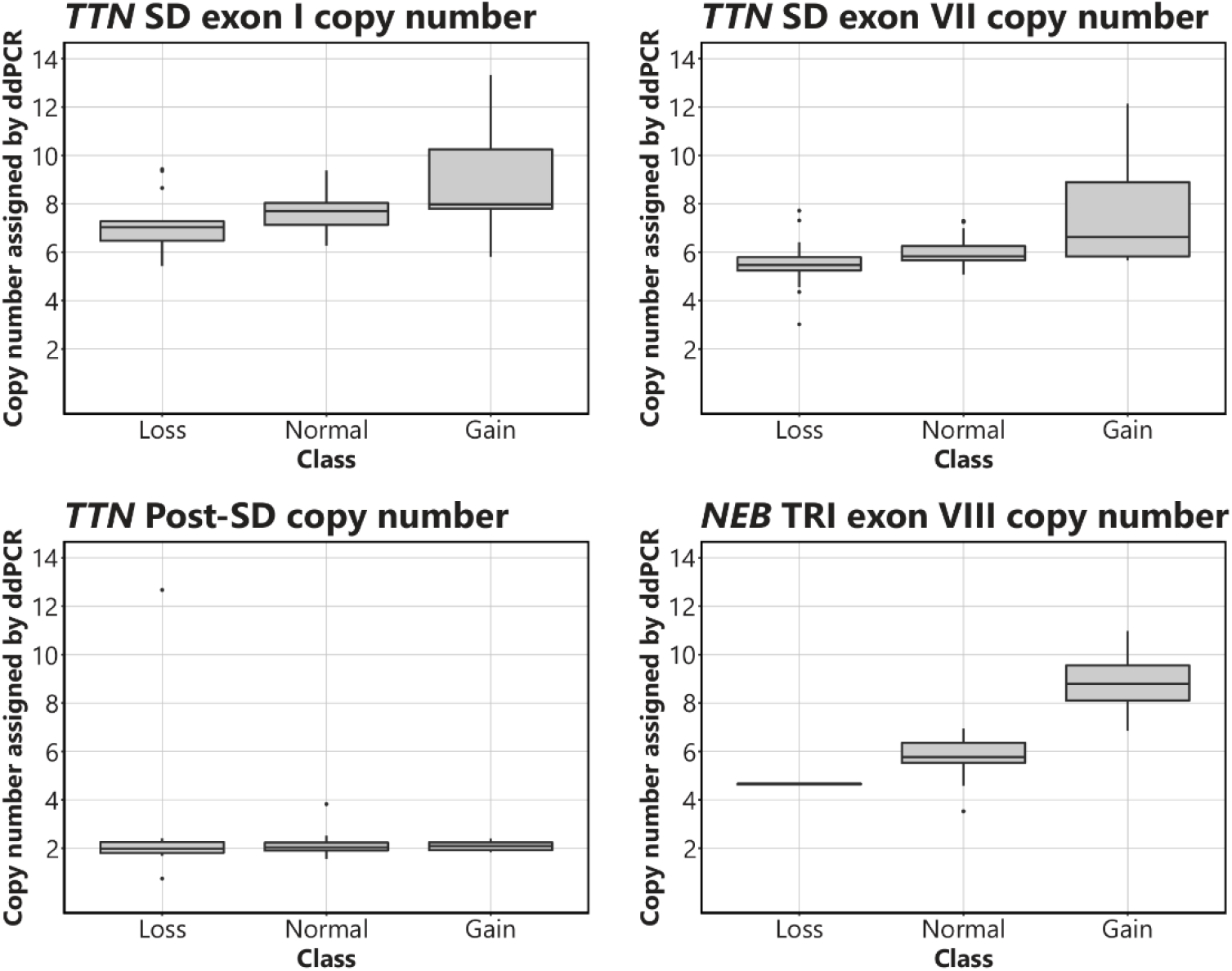
Boxplots visualizing the CN of the *TTN* SD exon I, *TTN* SD exon VII, *TTN* Post-SD and *NEB* TRI exon VIII assays in relation to the CN assigned to each targeted region by aCGH. The normal CNs for the assays are 8 for *TTN* SD exon I, 6 for *TTN* SD exon VII, 2 for *TTN* Post-SD, and 6 for *NEB* TRI exon VIII.

The mean accepted droplet count was 15,0042 (σ = 2,122.0, %CV = 14.1) for the *NEB* TRI exon VIII assay, 15,644 (σ = 2,159.9, %CV = 13.8) for the *TTN* SD exon I assay, 15,501 (σ = 2,322.3, %CV = 15.0) the *TTN* SD exon VII assay and 15,580 (σ = 2,482.4, %CV = 15.9) for the *TTN* Post-SD assay. The range, mean, standard deviation, and %CV values for accepted droplet numbers, target, and reference copies per 20 µl are presented in Supplemental Table 3. The range, mean, standard deviation, and %CV values for the ddPCR and aCGH estimated CNs for each ddPCR targeted sequence are presented in Supplemental Table 4.

#### One-way ANOVA and Tukey’s HSD

The one-way ANOVA indicated statistically significant differences between the ddPCR results for the different groups in each assay, except for the *TTN* Post-SD assay. The complete ANOVA and Tukey’s HSD test results are presented in Supplemental Table 5**Error! Reference source not found**.. Tukey’s HSD test for multiple comparisons found that the mean value of the ddPCR results was significantly different between normal and gain, and loss and gain samples in all assays except for the *TTN* Post-SD assay. No statistical significance could be found between normal and loss samples in any of the assays.

#### Pearson correlation

The Pearson correlation coefficient was 0.73 for the *TTN* SD exon I assay, 0.82 for the *TTN* SD exon VII assay, 0.13 for the *TTN* Post-SD assay, and 0.82 for the *NEB* TRI exon VIII assay, Additionally, we calculated a Pearson correlation coefficient to compare assays *TTN* SD exon I and *TTN* SD exon VII, yielding a value of 0.87.

#### Linear regression

The overall regression was statistically significant for the *TTN* SD exon I assay (adjusted R^2^ = 0.53, F(1,53) = 50.59, p < 0.00001), the *TTN* exon VII assay (adjusted R^2^ = 0.66, F(1,53) = 104.9, p < 0.00001), and the *NEB* TRI exon VIII assay (adjusted R^2^ = 0.67, F(1,53) = 113, p < 0.00001). The overall regression was not statistically significant for the *TTN* Post-SD assay (adjusted R^2^ = −0.002, F(1,53) = 0.89, p = 0.351). The complete unrounded results of the linear regression models are presented in Supplemental Table 6.

#### Intra-assay analysis

To assess the reproducibility of the assays, the intra-assay duplicate means, σ, and % CV were calculated along with an average %CV for all samples per assay. The intra-assay %CV mean was 4.40 for *TTN* SD exon I (n = 45), 3.97 for *TTN* SD exon VII (n = 42), 2.31 for *TTN* Post-SD (n = 40), and 3.77 for *NEB* TRI exon VIII (n = 42). The complete intra-assay analyses are presented in Supplemental Table 7.

#### Inter-assay analysis

To assess the repeatability of the assays, the inter-assay duplicate means, σ, and %CV were calculated along with an average %CV for all samples per assay. The inter-assay %CV mean was 12.75 for *TTN* SD exon I (n = 10), 3.95 for *TTN* SD exon VII (n = 13), 4.29 for *TTN* Post-SD (n = 15), and 3.51 for *NEB* TRI exon VIII (n = 12). The inter-assay analyses are presented in Supplemental Table 8.

## Discussion

Our novel ddPCR assays confirm the presence of recurring CNVs within the *TTN* SD region, as first seen on the NMD-CGH-array (18). CNVs in the *TTN* SD region is about three times more common than CNVs in the *NEB* TRI region. To our knowledge, this is the first time that CNVs within this region are acknowledged to this degree.

Our custom ddPCR assays for *TTN* SD exon I and *TTN* SD exon VII reliably recognise samples with gains in the *TTN* SD region, as supported by the statistical tests performed.

There was no statistical significance between the normal and loss groups in the *TTN* SD exon I and *TTN* SD exon VII assays. This is presumably due to two factors: the relatively low number of samples and thus the lack of power in the statistical methods, and the lack of CN amplitude variance within the deletion group. In the over 430 samples that we have previously analysed on our custom NM- and NMD-CGH-arrays (17, 18) we have yet to see losses of two or more copies of the *NEB* TRI region. We have, however, seen cases with up to 8 extra copies, yielding a total CN of 14 of the *NEB* TRI. It is also possible, that at least part of the suspected *TTN* SD region losses are methodological artefact due to the relatively low number of good quality aCGH probes that can be designed in the region. This, in turn, is limited by the repetitiveness and length of the *TTN* SD region.

The ddPCR method for the detection of CNVs within the *TTN* SD could theoretically be improved by designing more assays covering other sequences of the region. In our experience, however, in silico optimisation does not necessarily guarantee working assays. Furthermore, the exons within the repeating block of the *TTN* SD are highly similar, which limits the possibilities for unique assay design.

The intra- and inter-assay analyses showed adequate reproducibility and acceptable repeatability. The larger repeatability %CV values may be explained by differences in manual handling, such as differences in pipetting technique between the persons who produced the data. Both intra- and inter-assay analysis results could improve by using automated pipetting machinery.

The *TTN* SD is part of the PEVK-encoding region of titin. This region undergoes extensive tissue-specific alternative splicing giving rise to cardiac and skeletal muscle specific isoforms. The *TTN* SD exons seem to be missing from the cardiac isoforms (36). The PEVK region forms an elastic spring that modulates titin-based force in skeletal muscle through interactions with calcium and actin (37).

What remains to be elucidated is the exact CN of the *TTN* SD gains and losses. Furthermore, phenotype-genotype correlation analyses are needed to pinpoint a pathogenic threshold of the *TTN* CNVs. It is unlikely that a single repeat block gain or loss would significantly alter the phenotype, seen the size of the gene and its product. However, we hypothesize that a large enough size deviation of the PEVK region from normal may affect the elastic properties of titin and force generation in skeletal muscle.

Approximately 5% of the human genome consists of SD regions and are a significant feature of mammalian genomes (1-4). Repetitive regions, including SDs, are often neglected in both MPS-based and aCGH methods. While sequencing technology and analysis of sequence data have come a long way in terms of CNV detection (14, 15), the methodology has significant limitations regarding repetitive sequences. DNA amplification is a common step in MPS protocols, and in the case of repeated regions, it may severely distort the true repeat number in the subsequent analysis. More importantly, the length of the contigs that modern MPS technologies produce, is still insufficient to get a reliable alignment of these repetitive regions, which may span several megabases.

Attempts to sequence over the *NEB* TRI and *TTN* SD regions have so far been unsuccessful, as even the long-read sequencing methods often rely on an amplification step. As long-read sequencing methods develop in due time, we hope to sequence over both the *NEB* TRI and *TTN* SD regions.

## Data Availability

All relevant data in the present work are contained in the manuscript and its supporting information. Raw data produced in the present study are available upon reasonable request to the authors.

## Acknowledgements

We thank Marilotta Turunen for excellent technical assistance. We thank Erkka Valo for help with optimizing the R script. We thank Mari Ainola, Vesa-Petteri Kouri, and Soili Kytölä for their help in the set-up of the method, and the Musculoskeletal and Inflammation Research Group (TULES-Group) for providing access to the ddPCR machinery. We thank Carina Wallgren-Pettersson for her valuable comments on the manuscript.

## Author Contributions

Study concept and design: LS, VLL, KP, KK. Acquisition, analysis, and interpretation of data: LS. Drafting of the paper: LS. Critical revision of the paper for important intellectual content: VLL, KP, KK. Acquiring funding: KP, KK. Study supervision: VLL, KP, KK.

## Ethical approval

The study was approved by the Ethics Committee of the Children’s Hospital, University of Helsinki, Finland (approval number 6/E7/05). The approval was renewed by the Ethics Review Board of Helsinki University Hospital in 2021. Samples were obtained according to the declaration of Helsinki of 1975 and written consent was obtained from subjects, or data was analysed pseudonymously where appropriate.

## Competing interests

The authors declare no competing interests.

## Notes

### Competing Interest Statement

The authors have declared no competing interest.

### Funding Statement

This study was funded by Magnus Ehrnrooth foundation, Finska Laekaresaellskapet, Svenska Kulturfonden, Stiftelsen Dorothea Olivia, Karl Walter och Jarl Walter Perklens minne, Folkhaelsan Research Center.

### Author Declarations

The Ethics Committee of the Children's Hospital of the University of Helsinki, Finland gave ethical approval for this work (approval number 6/E7/05). The Ethics Review Board of the University of Helsinki, Finland, renewed the ethical approval in 2021. Samples were obtained according to the declaration of Helsinki of 1975 and written consent was obtained from subjects, or data was analysed pseudonymously where appropriate.

